# Intention to participate in a COVID-19 vaccine clinical trial and to get vaccinated against COVID-19 in France during the pandemic

**DOI:** 10.1101/2020.04.23.20076513

**Authors:** Maëlle Detoc, Sébastien Bruel, Paul Frappe, Elisabeth Botelho-Nevers, Amandine Gagneux-Brunon

## Abstract

**Background:** The world is facing the COVID-19 pandemic. Development of vaccine is challenging.

**Aim:** To determine the proportion of people who intend to get vaccinated against COVID-19 in France or to participate in a vaccine clinical trial.

**Methods:** We conducted an anonymous on-line survey from the 26th of March to the 20th of April 2020. Primary endpoints were the intention to get vaccinated against COVID-19 if a vaccine was available or participate in a vaccine clinical trial.

**Results:** Three thousand two hundred and fifty nine individuals answered the survey; women accounted for 67.4 % of the responders, 670 (20.6 %) were under 30 years of age, 1,502 (46.1 %) between 30-49 years, 803 (24.6 %) between 50-64 years, 271 (8.3%) between 65-80 years, 13 (0.4%) over 80 years of age. According to their statements, 2.512 participants (77.6%, 95 % CI 76.2-79 %) will certainly or probably agree to get vaccinated against COVID-19. Older age, male gender, fear about COVID-19, being healthcare workers and individual perceived risk were associated with COVID-19 vaccine acceptance Vaccine hesitancy was associated with a decrease in COVID-19 vaccine acceptance. One thousand and five hundred and fifty responders (47.6 % 95 % CI 45.9-49.3 %) will certainly or probably agree to participate in a COVID-19 vaccine clinical trial.

**Conclusions and Relevance:** Nearly 75 % and 48 % of the survey responders were likely to accept vaccination or participation in a clinical trial against COVID-19. Vaccine hesitancy will be the major barrier to COVID-19 vaccine uptake.

---

In December 2019, China reported the first cases of Coronavirus Disease (COVID-19) due to a new Coronavirus: the Severe Acute Respiratory Syndrome-Coronavirus 2 (SARS-CoV 2). The world is now facing a pandemic, more than a million of people have become infected worldwide, and cases are described in more than 200 hundred countries and [1].

In 2019, the World Health Organization identified ten threats to global health [2]. Among these threats, vaccine hesitancy, the risk of a global influenza pandemic, and the risk of emergence of high-threat pathogens such as Middle-East Respiratory Syndrome and/or Severe Acute Respiratory Syndrome were identified. Since this statement, COVID-19 had emerged. Developing COVID-19 vaccines is a crucial challenge, and several candidates are currently under basic development [3]. Some of them will be tested in Phase I trials in the weeks to come [4]. The time it takes to develop a vaccine is estimated to be 1 or 1.5 years as different steps are necessary during the clinical development of a vaccine [5]. Recruitment of volunteers in a vaccine clinical trial is a real challenge [6], and some trials are stopped du to difficulties in recruitment. Vaccine hesitancy may also has an impact on recruitment in COVID- 19 vaccine clinical trial [7]. After its clinical development, a COVID-19 vaccine will also face the challenge of acceptance by the general population in a post-crisis context.

The impact of the current pandemic on the intention to participate in a COVID-19 vaccine clinical trial and on the intention to get vaccinated against COVID-19 vaccine is not obvious. This is a particular concern in France, which has been shown to be the leader-country of vaccine hesitancy [8]. The aim of this study was to evaluate the impact of the pandemics on the intention to get vaccinated against COVID-19.

## Methods

We conducted an anonymous online survey (Lime Survey) from the 26^th^ of March to the 12^th^ of March 2020 among adult general population and adult patients. The survey was proposed to individuals via social networks (Facebook, Twitter), shared by e-mail, on the website of the University Hospital of Saint-Etienne (France), and in centers for COVID-19 diagnosis and in medical centers. We developed a standardized questionnaire based on a literature review.

The questionnaire addressed: (1) demographical characteristics (age, chronic medical conditions), (2) fears about COVID-19, (3) history of vaccination against pandemic H1N1 influenza and seasonal influenza, (4) intention to get vaccinated if a COVID-19 vaccine was available, (5) vaccine hesitancy. We evaluated participants’ self-reported vaccine hesitancy according to the WHO definition (declining a vaccine considered dangerous or unnecessary; delaying a vaccine because of doubts about it, accepting a vaccine despite doubts about its efficacy or safety)[9]. If a participant answered yes to one of these proposals, he or she was considered to be “vaccine hesitant”.

The questionnaire included items to be answered on a 5-level Likert scale including a “don’t know” option to evaluate intention to participate in a clinical trial and to get vaccinated if a vaccine was available. We combined survey responses into two categories (strongly agree/agree, don’t know/disagree/strongly disagree) and ran ordinal regression models to examine demographic and attitudinal factors predictive of respondents’ willingness to get vaccinated against COVID-19. To identify suitable candidate variables for regression models, we first conducted univariate analysis using a chi-squared test. Candidates that were significant at P < 0.1 in univariate analyses were then included in a multivariable regression model. Data were analyzed using SPSS version 24.0.

The protocol complied with the data privacy laws of the National Commission for Informatics and Civil Liberties and was approved by the institutional review board with the number IRBN422020/CHUSTE.

## Results

During the study period, 3,656 people opened the web links for the on-line survey, 3,259 (89.1 %) people answered the on-line questionnaire. Demographic characteristics are displayed in table 1. Women accounted for 67.4 % of the responders. Seven hundred and eighty-seven (24.1%) responders reported a chronic medical conditions, 68 (2.1%) reported a history of diabetes mellitus, 260 (8.0%) of the study participants reported a history of hypertension, 92 (2.8%) a history of cardiac disease, 139 (4.3%) a history of chronic lung disease, and 63 (1.9%) were receiving an immunosuppressive medication. Vaccine hesitancy was observed in 1,150 responders (35.3% 95% CI 33.6-36.9%) were considered as vaccine hesitant.

**Table 1:** Demographic characteristics of the responders to the on-line survey, data are expressed by n and (%)

|  | N=3,259 (%) |
| --- | --- |
| Gender |  |
| Male | 1,063 (32.6) |
| Female | 2,196 (67.4) |
| Age |  |
| Under 30 years old | 670 (20.6) |
| 30-49 years old | 1,502 (46.1) |
| 50-64 years old | 803 (24.6) |
| 65-80 years old | 271 (8.3) |
| Over 80 years old | 13 (0.4) |
| Health care workers |  |
| Yes | 1,421 (43.6) |
| No | 1,838 (56.4) |
| Chronic medical conditions | 627 (19.2) |
| Hypertension | 260 (8.0) |
| Diabetes | 68 (2.1) |
| Chronic lung disease | 139 (4.3) |
| Chronic cardiac failure | 92 (2.8) |
| Chronic renal insufficiency | 2 (0.1) |
| Immunosuppressive medication | 63 (1.9) |
| Fear about COVID-19 |  |
| Yes, a lot | 766 (23.5) |
| Yes, a little | 1,668 (51.2) |
| I don't know | 176 (5.4) |
| Not really | 549 (16.8) |
| No | 100 (3.1) |
| Perceived individual risk to catch COVID-19 |  |
| Yes, a lot | 644 (19.8) |
| Yes, a little | 1,480 (45.4) |
| I don't know | 426 (13.1) |
| Not really | 615 (18.9) |
| No | 94 (2.9) |
| Vaccine hesitancy |  |
| Have you ever refused a vaccine for yourself or a child because you considered it as useless or dangerous? | 395 (12.1) |
| Have you ever postponed a vaccine recommended by a physician? | 341 (10.5) |
| Have you ever had a vaccine for a child or yourself despite doubts about its efficacy? | 570 (17.5) |
| Global vaccine hesitancy | 1,150 (35.3) |
| Acceptance to participate in a COVID-19 vaccine clinical trial | 1,552 (47.6) |
| Willingness to get vaccinated against COVID-19 if a vaccine was available | 2,530 (77.6) |

Two thousand four hundred-thirty four (74.7% 95 % CI 73.2-76.2%) responders had fears about COVID-19; 2,124 (65.2% 95% CI 63.6-66.8%) responders considered themselves at risk for COVID- 19. Nine hundred and seventy-two responders (29.8% 95% CI 28.2-31.3 %) declared that they got vaccinated against H1N1 pandemic influenza, in 2009.

### Willingness to get vaccinated against COVID-19

According to their statements, 2,512 participants (77.6%, 95 % CI 76.2-79.0 %) will certainly or probably be willing to get vaccinated against COVID-19. Among the 1,063 men, 883 (83.1% 95% CI 80.8-85.3%) are COVID-19 vaccine acceptors, 1,629 women among the 2,196 responders (74.2 % 95 % CI 72.3-76.0%) are COVID-19 vaccine acceptors (p<0.005). The proportion of vaccine hesitant responders who would probably be willing to get vaccinated against COVID-19 vaccine was 61.9 % (95 % CI 59.1-64.7 %) during the current pandemics. The proportion of healthcare workers willing to get vaccinated was 81.5 %, and this proportion was 73.7 % in non-healthcare workers (p<0.005). Factors associated with COVID-19 acceptance are displayed in Table 2. In multivariable analysis, older age, male gender, fear about COVID-19, be healthcare workers and individual perceived risk remained associated with COVID-19 vaccine acceptance. Vaccine hesitancy was associated with lower acceptance of a COVID-19 vaccine.

**Table 2.** Factors associated with COVID-19 vaccine acceptance expressed with odds ratio (ref: reference* p<0.1, ** p<0.005), in multivariable analysis, only variables with a p-value<0.1 in univariate analysis were integrated in the model.

|  | Univariate analysis | Multivariable analysis |
| --- | --- | --- |
| **Gender** |  |  |
| Female | Ref | Ref |
| Male | 1.707 (1.417-2.058)** | 1.878 (1.529-2.306)** |
| **Age** |  |  |
| Under 30 years old | Ref | Ref |
| 30-49 years old | 1.506 (1.231-1.842)** | 1.532 (1.230-1.909)** |
| 50-64 years old | 2.246 (1.758-2.870)** | 1.445 (1.259-1.658) ** |
| 65-80 years old | 2.692 (1.854-3.908)** | 1.395 (1.198-1.624) ** |
| Over 80 years old | 5.593 (0.723-43.293)* | 1.412 (0.827-2.412) |
| **Healthcare workers** |  |  |
| No | Ref | Ref |
| Yes | 1.574 (1.329-1.864)** | 1.533 (1.269-1.851)** |
| **Chronic medical conditions** |  |  |
| No | Ref |  |
| Yes | 1.196 (0.983-1.455)* | 0.891 (0.713-1.113) |
| **Fear about COVID-19** |  |  |
| No | Ref | Ref |
| Yes | 2.095 (1.757-2.499)** | 2.445 (1.998-2.991)** |
| **Perceived individual risk** |  |  |
| No | Ref | Ref |
| Yes | 1.831 (1.549-2.163)** | 1.510 (1.269-1.851)** |
| **Vaccine hesitancy** |  |  |
| No | Ref | Ref |
| Yes | 0.279 (0.236-0.331)** | 0.275 (0.230-0.329)** |

### Willingness to participate in a COVID-19 vaccine clinical trial

One thousand and five hundred and fifty two responders (47.6 % 95 % CI 45.9-49.3 %) will certainly or probably be willing to participate in a COVID-19 vaccine clinical trial. Among the 1.063 men, 634 (59.6 % 95 % CI 56.7-62.5 %) will probably accept to participate in a COVID-19 vaccine clinical trial, this proportion is significantly greater than women (41.8 % 95 % CI 39.7-43.9 %, p< 0.005). The percentage of potential participants in a COVID-19 vaccine clinical trial was 56.8 % (53.4-60.2 %) in the 50-64 years age group, and 58.7 % (95 % CI 52.8-64.6 %) in the 65-80 years age group. Healthcare workers are more prone to participate in a vaccine clinical trial than non-healthcare workers (50.5 % vs 45.4 %, p<0.005). Factors associated with COVID-19 acceptance are displayed in Table 3.

**Table 3.** Factors associated with potential participation in a COVID-19 vaccine trial expressed with odds ratio (* p<0.1, ** p<0.005), in multivariable analysis, only variables with a p-value<0.1 in univariate analysis were integrated in the model.

|  | Univariate analysis | Mutivariable analysis |
| --- | --- | --- |
| **Gender** |  |  |
| Male | 2.057 (1.773-2.388)** | 1.881 (1.532-2.309)** |
| Female | Ref | Ref |
| **Age** |  |  |
| Under 30 years old | Ref | Ref |
| 30-49 years old | 1.043 (0.868-1.254) | 1.518 (1.220-1.888)** |
| 50-64 years old | 1.797 (1.461-2.211)** | 1.481 ( 1.297-1.690)** |
| 65-80 years old | 1.941 (1.458-2.585)** | 1.408 (1.229-1.612)** |
| Over 80 years old | 0.608 (0.185-1.993) | 1.494 (0.881-2.531) |
| **Healthcare workers** |  |  |
| No | Ref | Ref |
| Yes | 1.223 (1.065-1.405)** | 1.540 (1.275-1.860)** |
| **Chronic medical conditions** |  |  |
| No | Ref |  |
| Yes | 1.085 (0.924-1.274) |  |
| **Fear about COVID-19** |  |  |
| No | Ref | Ref |
| Yes | 0.895 (0.764-1.048) | 0.928 (0784-1.099) |
| **Perceived individual risk** |  |  |
| No | Ref | Ref |
| Yes | 1.226 (1.061-1.417)** | 1.235 (1.061-1.439)** |
| **Vaccine hesitancy** |  |  |
| No | Ref | Ref |
| Yes | 0.502 (0.433-0.582)** | 0.512 (0.4411-0.595)** |

## Discussion

In this on-line survey, we observed that nearly 77 % of the responders would accept a vaccine against COVID-19 although 35 % of the responders were qualified as “vaccine hesitant”. Moreover, 47.6 % of the responders would accept to participate in a clinical trial for a COVID-19 vaccine. Concerning the intention to get vaccinated, our results are similar to the results of the longitudinal Coconel study conducted by the Observatoire Régional de Santé Provence Alpes Côte d’Azur [10]. Similar to their results, we observed that men were more prone to get vaccinated than women. Women accounted for the vast majority of our study responders, suggesting that in real-life settings, COVID-19 vaccine acceptance could be greater. In addition, older individuals are more prone to get vaccinated in both studies, this is probably due to a greater perceived risk of getting infected and developing a severe disease in older people. In the same vein, healthcare workers were more prone to get vaccinated or to participate in a vaccine clinical trial than the others, it is probably due, in part, to a greater perceived risk to get infected. Healthcare workers are particularly vulnerable and accounted for 10 % of the infected people in Italy [11]. On the contrary, expecting a low infection risk is associated with a lower willingness to get vaccinated [12].

Around the half of the responders will accept to participate in a COVID-19 vaccine clinical trial. In the context of a clinical trial, men were also more prone to participate. Fears about COVID-19 were not associated with willingness to participate in a clinical trial. However, individuals who considered themselves at-risk for COVID-19 infection were more prone to accept to participate in a clinical trial for a vaccine. Out of the context of the pandemics, the feeling to be at-risk for a disease was not the main motivation to participate in a vaccine clinical trial [7]. Moreover, older age was associated (except in the group over 80 years) with willingness to participate in a COVID-19 vaccine clinical trial, this observation contrast with a previous study about willingness to participate in a vaccine clinical trial [7]. This observation suggests that in the pandemics context, individuals are more prone to participate in a clinical trial for a vaccine.

The proportion of French People favorable to a COVID-19 vaccine can appear low, but we have to put this in perspective with the fact that the vaccine would be a new vaccine, and that vaccine coverage against 2009 H1N1 pandemic influenza was only 11.1 % in France [13]. However, a greater proportion of responders to our survey declared they had been vaccinated against 2009 H1N1 pandemic influenza, so this observation may suggest that the responders are more pro-vaccine than the general population in France, and more often healthcare workers. We observed a similar prevalence of vaccine hesitancy than in a recent study in France, which identified a prevalence of 40 % [9]. One limitation of our work may be the use of social media to recruit study participants. Social media users who use these medias as health information sources are more prone to get vaccinated against seasonal influenza in the United States of America [14]. Furthermore, a great number of healthcare workers (43.6 % of the responders) answered the survey and we observed that healthcare workers were more prone to get vaccinated or to participate in a vaccine clinical trial independently of the perceived risk to get contaminated. However, vaccine hesitancy also affects healthcare workers [15–17]. In our study sample, vaccine hesitancy affects 29.3 % of the healthcare workers and 39.9 % of the non-healthcare workers. Our sample is not completely representative of the French general population. In addition, we did not precise in the survey, the type of clinical trials, and intention to participate may change between early and later phases clinical trials [7].

In conclusion, during the pandemics, around 75 % of the French people would agree to get vaccinated. Due to the burden of the disease, and the potential natural immunity, it may well be possible that this proportion would be enough to obtain a herd effect [18]. Vaccine hesitancy is the major barrier to implement vaccines campaign even in a context of a pandemic. Around fifty percent will agree to participate in a COVID-19 vaccine clinical trial, we can hope that vaccine trials would not be stopped because of recruitment difficulties.

## Data Availability

Data are available after request to the corresponding author

## Acknowledgment

Pr Glyn Thoiron for English editing

Co-authors have no conflict of interest to declare for this work.

## References

1. Coronavirus disease (COVID-19) Situation Dashboard [Internet]. [accessed 3 apr. 2020]. Available: https://experience.arcgis.com/experience/685d0ace521648f8a5beeeee1b9125cd

2. Ten health issues WHO will tackle this year [Internet]. [accessed 3 apr 2020]. Available: https://www.who.int/news-room/feature-stories/ten-threats-to-global-health-in-2019

3. Lurie N, Saville M, Hatchett R, Halton J. Developing Covid-19 Vaccines at Pandemic Speed. New England Journal of Medicine. 30 march 2020;0(0)

4. Le TT, Andreadakis Z, Kumar A, Román RG, Tollefsen S, Saville M,et al. The COVID-19 vaccine development landscape. Nature Reviews Drug Discovery [Internet]. 9 apr 2020 [accessed 12 apr 2020]; Disponible sur: http://www.nature.com/articles/d41573-020-00073-5

5. Eyal N, Lipsitch M, Smith PG. Human challenge studies to accelerate coronavirus vaccine licensure. J Infect Dis [Internet]. [accessed 3 apr 2020]; Available on: http://academic.oup.com/jid/advance-article/doi/10.1093/infdis/jiaa152/5814216

6. Cobb EM, Singer DC, Davis MM. Public interest in medical research participation: differences by volunteer status and study type. Clin Transl Sci. 2014;7(2):145–9.

7. Detoc M, Launay O, Dualé C, Mutter C, Le Huec J-C, Lenzi N, et al. Barriers and motivations for participation in preventive vaccine clinical trials: Experience of 5 clinical research sites. Vaccine. 2019;37(44):6633–9.

8. Larson HJ, de Figueiredo A, Xiahong Z, Schulz WS, Verger P, Johnston IG, et al. The State of Vaccine Confidence 2016: Global Insights Through a 67-Country Survey. EBioMedicine. 2016;12:295–301.

9. Rey D, Fressard L, Cortaredona S, Bocquier A, Gautier A, Peretti-Watel P, et al. Vaccine hesitancy in the French population in 2016, and its association with vaccine uptake and perceived vaccine risk–benefit balance. Eurosurveillance. 2018;23(17):17–00816.

10. Projets de recherche | ORS Paca [Internet]. [Accessed 13 avr 2020]. Available on: http://www.orspaca.org/covid19/projets-recherche

11. Chirico F, Nucera G, Magnavita N. COVID-19: Protecting Healthcare Workers is a priority. Infect Control Hosp Epidemiol. 2020;1–4.

12. Determann D, de Bekker-Grob EW, French J, Voeten HA, Richardus JH, Das E, et al. Future pandemics and vaccination: Public opinion and attitudes across three European countries. Vaccine. 2016;34(6):803–8.

13. Guthmann J-P, Fonteneau L, Bonmarin I, Lévy-Bruhl D. Influenza vaccination coverage one year after the A(H1N1) influenza pandemic, France, 2010-2011. Vaccine. 2012;30(6):995–7.

14. Ahmed N, Quinn SC, Hancock GR, Freimuth VS, Jamison A. Social media use and influenza vaccine uptake among White and African American adults. Vaccine. 2018;36(49):7556–61.

15. Killian M, Detoc M, Berthelot P, Charles R, Gagneux-Brunon A, Lucht F, et al. Vaccine hesitancy among general practitioners: evaluation and comparison of their immunisation practice for themselves, their patients and their children. Eur J Clin Microbiol Infect Dis. 2016;35(11):1837–43.

16. Agrinier N, Le Maréchal M, Fressard L, Verger P, Pulcini C. Discrepancies between general practitioners’ vaccination recommendations for their patients and practices for their children. Clinical Microbiology and Infection. 2017;23(5):311–7.

17. Wilson R, Zaytseva A, Bocquier A, Nokri A, Fressard L, Chamboredon P, et al. Vaccine hesitancy and self-vaccination behaviors among nurses in southeastern France. Vaccine. 2020;38(5):1144–51.

18. John TJ, Samuel R. Herd immunity and herd effect: new insights and definitions. Eur J Epidemiol. 2000;16(7):601–6.

